# Cardiovascular Resilience in Familial Hypercholesterolemia: Genomic Signatures from a Founder Population Highlight *IL34* as a Candidate Gene Associated with Event-Free Survival

**DOI:** 10.64898/2026.07.08.26357597

**Authors:** Etienne Khoury, Miriam Larouche, Alex Lauzière, Iulia Iatan, Diane Brisson, Daniel Gaudet

**Affiliations:** Department of Medicine, Université de Montréal and ECOGENE-21, Canada; Lipid Clinic and service of Cardiology, Chicoutimi Hospital, Canada; Lipidology, metabolism and internal medicine clinic, Chicoutimi, Canada; Research Institute of the McGill University Health Centre, Montreal, Quebec, Canada

**Author notes:** **Corresponding author and email address:** Daniel Gaudet, 930 Jacques Cartier Est, Room B-210, Chicoutimi, Canada, G7H 7K9.

**Keywords:** Interleukin-34, Familial Hypercholesterolemia, Cardioprotective Variant, Survival, whole-exome sequencing, Cardiovascular risk prevention

## Abstract

**Background:** Familial hypercholesterolemia (FH) is a semi-dominant genetic disorder characterized by lifelong elevation of low-density lipoprotein cholesterol (LDL-C) and a markedly increased risk of premature atherosclerotic cardiovascular disease (CVD). Despite this elevated risk, some individuals with FH survive beyond 70 years of age without developing clinical CVD. This study aimed to identify genetic variants associated with protection against cardiovascular events and to uncover mechanisms contributing to this resilience phenotype.

**Methods:** Whole-exome sequencing (WES) was performed in 243 French-Canadian heterozygous FH individuals carrying the pathogenic *LDLR* c.259T>G (p.Trp87Gly) variant. After stratification by age and cardiovascular event (CVE) status, 35 individuals with premature CVE and 20 individuals aged ≥70 years who remained free of CVE despite spending several decades in the pre-statin era were selected for comparative analysis.

**Results:** Variant annotation and quality-control validation using Firth logistic regression identified 12 genetic variants potentially associated with cardiovascular resilience. Among these, a stop-gain variant resulting from the single-nucleotide polymorphism rs4985556 in *IL34* demonstrated the strongest association with event-free survival (allele frequency in CVE− = 0.25 vs. CVE+ = 0.00; χ² = 19.25; *P* = 1.15 × 10⁻⁵). The *IL34* stop-gain variant (c.639C>A [p.Tyr213*]) was associated with a markedly increased likelihood of cardiovascular event-free survival (OR = 20.9; 95% CI, 2.7–2841.7; *P* = 3.45 × 10⁻⁴).

**Conclusion:** These findings identify *IL34* as a potential cardiovascular resilience gene and highlight novel genetic determinants that may protect against cardiovascular events despite lifelong exposure to elevated LDL-C levels. In addition to *IL34*, eleven variants showed strong associations with a CVE-free phenotype and warrant further investigation to elucidate their biological mechanisms and potential relevance for cardiovascular disease prevention.

## Introduction

Familial hypercholesterolemia (FH) is an autosomal semi-dominant disorder characterized by prolonged exposure to elevated plasma levels of low-density lipoprotein cholesterol (LDL-C), premature atherosclerosis, and xanthomatosis (1, 2). FH is primarily associated with pathogenic variants in three genes: *LDLR* (low-density lipoprotein receptor), *APOB* (apolipoprotein B), and *PCSK9* (proprotein convertase subtilisin/kexin type 9). Less frequently, variants in *LDLRAP1* (low-density lipoprotein receptor adaptor protein 1), *APOE* (apolipoprotein E), and *LIPA* (lysosomal acid lipase) have also been implicated (3–5).

FH affects approximately 1 in 200–300 individuals worldwide (1, 6). However, its prevalence is substantially higher in the French-Canadian population and is particularly elevated in Eastern Quebec, where it may affect as many as 1 in 120 individuals (7). More than 60% of FH cases in this region are attributable to two *LDLR* mutations: a >15 kb deletion (class 1 mutation) involving the promoter and exon 1, and the missense variant p.Trp87Gly (rs121908025) located in exon 3 (class 3 mutation). A panel of seven mutations accounts for the majority of FH cases in this founder population (7).

Heterozygous carriers of the p.Trp87Gly variant are at high risk of premature coronary artery disease (CAD) if left untreated (8–11). In addition, homozygous FH (HoFH) individuals carrying this variant have been identified in this population. In the absence of treatment, these patients present a severe phenotype characterized by early-onset CAD and markedly reduced life expectancy (12–15). Despite maximally tolerated statin therapy and/or combination treatment with ezetimibe and PCSK9 inhibitors, many patients with FH fail to achieve recommended LDL-C targets. Moreover, a significant proportion of patients are unable to tolerate high-intensity statin therapy because of muscular or non-muscular adverse effects (16, 17).

Founder populations provide unique opportunities to identify genetic variants that exert strong effects on disease expression, as certain alleles may be enriched relative to those observed in more genetically diverse populations (18, 19). Some of these variants may increase disease susceptibility, whereas others may confer protection against specific diseases. Several untreated or suboptimally treated FH patients from the French-Canadian cohort included in our previous studies have remained free of cardiovascular events (CVE) beyond the age of 70 years, with some HoFH individuals surviving beyond 70 years of age despite experiencing cardiovascular events. This observation is particularly remarkable given the lifelong exposure to markedly elevated LDL-C levels associated with FH, the suboptimal treatment status of many patients, and the relatively recent availability of highly effective lipid-lowering therapies.

We therefore hypothesized that a subset of patients with HeFH or HoFH may carry genetic factors that confer protection against atherosclerotic CVE and/or promote long-term survival despite lifelong exposure to elevated LDL-C levels and high expected atherosclerotic cardiovascular disease (ASCVD) risk. We previously investigated biological and environmental factors associated with this unexpected survival phenotype and identified female sex, higher HDL-C levels, elevated adiponectin concentrations and non-smoking status associated with CVE-free survival (20). The present study extends this work by identifying genomic determinants that may also contribute to cardiovascular resilience among patients aged ≥70 years carrying FH-causing *LDLR* variants. Identifying such resilience-associated variants may provide novel insights into the mechanisms underlying cardiovascular resilience and uncover potential therapeutic targets for preventing cardiovascular disease or mitigating its clinical consequences.

## Methods

### Subjects

A total of 243 French-Canadian FH subjects heterozygous for the pathogenic *LDLR* variant c.259T>G (p.Trp87Gly) were included in this study. The presence of the *LDLR* p.Trp87Gly mutation was determined using PCR-restriction fragment length polymorphism (PCR-RFLP) analysis. Subjects were subsequently stratified according to age and the occurrence of CVE requiring medical care, including myocardial infarction, unstable angina, coronary artery bypass grafting, and percutaneous transluminal coronary angioplasty. Two groups were defined: Group 1 comprised subjects aged ≥70 years (mean age: 78.0 ± 6.5) with no history of CVE [CVE(-)], whereas Group 2 included subjects aged <70 years (mean age: 61.0 ± 6.0) with documented CVE [CVE(+)] (Figure 1). Notably, individuals in the CVE(-) group had spent approximately 30 to 50 years of their lives in the pre-statin era or before statin therapy became widely adopted following the publication of the Scandinavian Simvastatin Survival Study (4S) at the end of 1994 (21). Consequently, these subjects experienced prolonged exposure to elevated LDL-C levels in the absence of effective lipid-lowering treatment. All clinical features, including type 2 diabetes (T2D), hypertension, and body mass index (BMI), as well as the lipid profile and adiponectin levels were collected as previously described (20). Subjects were screened at the Chicoutimi Hospital Lipid Clinic or ECOGENE-21 Clinical Research Center and agreed to participate in studies on genetic determinants of T2D, CAD and dyslipidemia. This project is a sub-analysis of the “Study of genetic determinants of diabetes, CAD and dyslipidemia in the French-Canadian population”, which has received the approval of either the Chicoutimi Hospital Ethics Committee or Advarra (IRB Services) (#MOD00390737), in accordance with the Declaration of Helsinki. Subjects gave their informed consent to participate in this study and were assigned a code that systematically de-identifies all clinical data. Subjects were selected to be included in the present study based on the availability of data.

**Figure 1.**
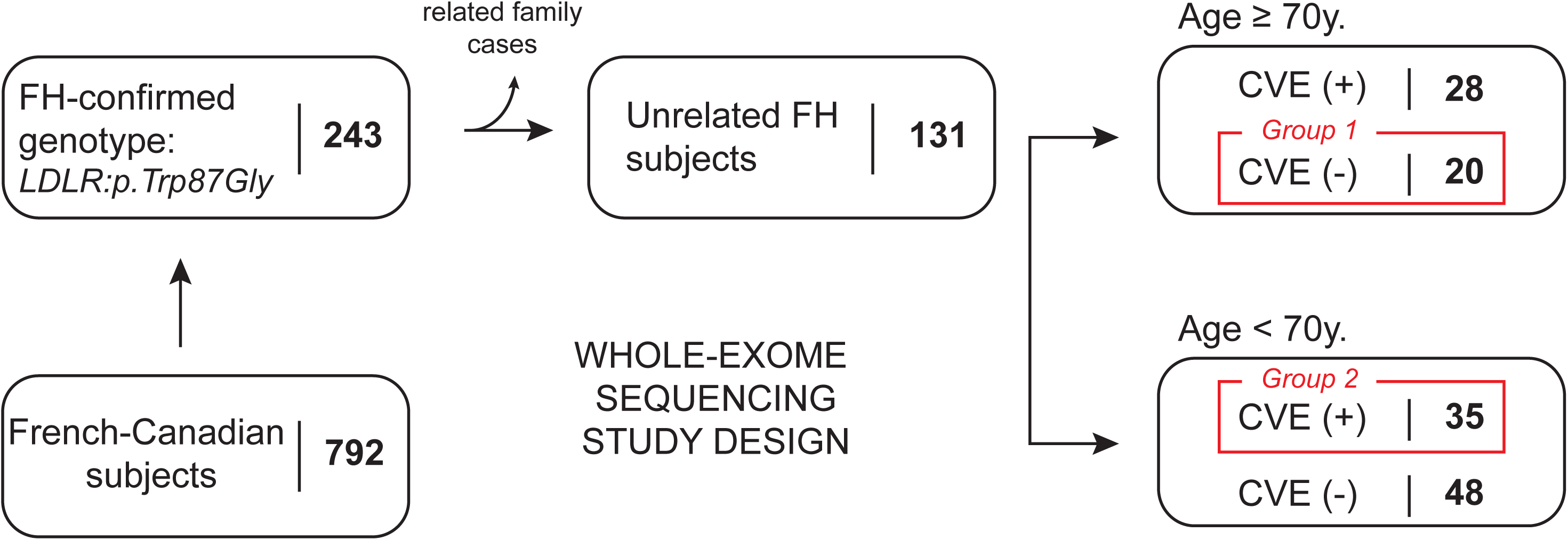
Study design. A total of 243 French-Canadian patients with genetically confirmed heterozygous FH carrying the *LDLR* c.259T>G (p.Trp87Gly) variant were selected from an initial cohort of 792 individuals. Following the exclusion of related individuals (one proband retained per pedigree), subjects were stratified according to age (<70 years versus ≥70 years) and history of documented cardiovascular events (CVE(+) versus CVE(-), respectively). The red boxes indicate the subjects selected for the primary analysis aimed at identifying genetic variants associated with cardiovascular event-free survival.

### Blood Samples

Blood samples were collected after a 12-hour overnight fast. Total cholesterol and triglyceride (TG) concentrations were measured using enzymatic assays. LDL-C levels were calculated using the Friedewald formula or measured directly when TG concentrations exceeded 4.5 mmol/L. Ultracentrifugation was performed to determine the cholesterol content of LDL and high-density lipoprotein (HDL) particles. Apolipoprotein B (ApoB) and apolipoprotein A-I (ApoA-I) concentrations were measured by nephelometry. Lipoprotein(a) [Lp(a)] levels were determined using enzyme-linked immunosorbent assays (ELISA) (B-Bridge International, Inc., San Jose, CA, USA).

### Whole-Exome Sequencing

Whole-exome sequencing (WES) was performed on high-quality genomic DNA (∼1 μg). Genomic DNA was extracted from whole blood using the QIAamp DNA Blood Kit (Qiagen, Hilden, Germany). Exome libraries were prepared using the VCRome v2.1 capture platform (NimbleGen, Pleasanton, CA, USA). Captured libraries were sequenced on the Illumina HiSeq 2500 platform (Illumina, San Diego, CA, USA) using 2 × 150 bp paired-end sequencing chemistry. Paired-end reads were aligned to the human reference genome (GRCh37/hg19) using the Burrows–Wheeler Aligner MEM algorithm (BWA-MEM) (22). The Genome Analysis Toolkit (GATK v3.1) was used for local realignment around insertion/deletion (indel) sites and for variant calling using the HaplotypeCaller module.

### Annotation and Quality Control

Following variant calling of single nucleotide polymorphisms (SNPs) and insertion/deletion variants (indels), variant call format (VCF) files were annotated using reference SNP cluster identifiers (rsIDs), variant consequences predicted by the Ensembl Variant Effect Predictor (VEP), and functional annotations based on Gene Ontology (GO) molecular function terms. Quality control procedures included the exclusion of variants with >2% missing genotypes and variants with a minor allele frequency (MAF) <1%. Variants were subsequently restricted to coding and exonic regions and further annotated according to their predicted functional consequences, including synonymous, missense, nonsense, and frameshift variants. Additional filtering was performed to prioritize variants with a high predicted impact on gene function, protein expression, and biological processes.

### Association Testing

To identify genetic markers associated with cardiovascular event-free (CVE-free) survival and resistance to atherosclerotic CVE among elderly FH patients, WES data were analyzed according to both age (<70 years vs. ≥70 years) and reported CVE status [CVE(+) vs.CVE(-)] (Figure 1).

Genome-wide association results were visualized using Manhattan plots generated in R version 3.4.1 with the *qqman* package (Figure S1). Although studies of this size are generally underpowered for genome-wide association analyses at conventional genome-wide significance thresholds (*P* < 5 × 10⁻⁸), we anticipated that the reduced genetic heterogeneity of this founder FH cohort, combined with the exclusion of related individuals, would facilitate the identification of variants with relatively large effects. Therefore, an exploratory significance threshold of *P* < 1 × 10⁻³ was used to prioritize candidate variants for subsequent validation analyses. Given the exploratory and hypothesis-generating nature of this study, no formal correction for multiple testing was applied, and the identified variants should therefore be interpreted as candidate loci requiring independent validation. Variants showing the strongest associations were subsequently selected for internal statistical confirmation using Firth logistic regression analysis, a method particularly well suited for sparse datasets and low-frequency variants. This approach was used to estimate the relative contribution of each variant to CVE-free survival among FH subjects aged ≥70 years, expressed as odds ratios (ORs) and corresponding 95% confidence intervals (95% CIs).

### Statistical analysis

Based on the initial study sample, statistical analyses of lipid profile variables were performed by comparing CVE-free survivors [CVE(-)] with younger subjects who had experienced premature CVE [CVE(+)]. Categorical variables were compared using Pearson’s χ² test, which was also used to compare allele frequencies (AFs) between the CVE(-) and CVE(+) groups. Variables exhibiting a skewed distribution were log₁₀-transformed prior to statistical analysis. Given the relatively small sample size and the low frequencies of several genetic variants, Firth logistic regression models were fitted to obtain more reliable parameter estimates and to assess the association between candidate variants and CVE-free survival. The strength of these associations was expressed as ORs with corresponding 95% confidence intervals (95% CIs). All statistical tests were two-sided, and statistical significance was defined as *P* < 0.05. Statistical analyses were performed using SPSS software (version 21.0; IBM SPSS Statistics, Chicago, IL, USA).

## Results

Among HeFH subjects carrying the *LDLR* c.259T>G (p.Trp87Gly) variant, women were significantly more prevalent in the CVE(-) group (≥70 years) than in the CVE(+) group (<70 years) (75% vs. 26%, *P* = 0.0004) (Table 1). The premature CVE(+) group exhibited significantly lower HDL-C, ApoA-I as well as adiponectin concentrations compared to the CVE(-) group. No significant differences were observed for other lipid-related parameters, including LDL-C and ApoB concentrations (Table 1).

**Table 1.** Demographics for age-stratified cardiovascular event and event-free patients with *LDLR*:c.259T>G (p.Trp87Gly) HeFH.

|  | **FH CVE (+) <70y.**<br>(N=35) | **FH CVE (-) ≥70y.**<br>(N=20) | **P-value** |
| --- | --- | --- | --- |
| Female (%) | 25.7 | 75.0 | **0.0004** |
| Age (years) | 61.0 ± 6.0 | 78.0 ± 6.5 | **P <0.001\*** |
| Age of 1st CVE (years) | 39.6 ± 1.2 | - | - |
| Type 2 Diabetes (%) | 8.6 | 10 | 0.9 |
| Hypertension (%) | 25.7 | 35 | 0.5 |
| Body Mass Index (BMI) | 25.9 ± 3.48 | 28.17 ± 6.88 | 0.09 |
| Total cholesterol (mmol/L) | 8.8 ± 1.7 | 9.5 ± 1.6 | 0.2 |
| HDL-cholesterol (mmol/L) | 1.06 ± 0.27 | 1.27 ± 0.30 | **0.01\*** |
| LDL-cholesterol (mmol/L) | 7.1 ± 1.5 | 7.5 ± 1.4 | 0.3 |
| Non-HDL-cholesterol (mmol/L) | 7.8 ± 1.7 | 8.2 ± 1.6 | 0.4 |
| Total-C/HDL-C | 9.1 ± 3.5 | 7.8 ± 2.1 | 0.2 |
| Triglycerides (mmol/L) | 1.3 [1.0-1.9] | 1.6 [1.2-2.5] | 0.1 |
| Apolipoprotein A-I (g/L) | 1.21 ± 0.29 | 1.52 ± 0.25 | **0.009\*** |
| Apolipoprotein B (g/L) | 1.38 ± 0.43 | 1.44 ± 0.41 | 0.7 |
| Lipoprotein(a) (mg/dl) | 45.7 [10.7-66.2] | 12.4 [5.4-47.5] | 0.05 |
| Adiponectin (mg/mL) | 5.44 [4.12-9.08] | 8.88 ± [6.82-10.83] | **0.044\*** |

Numerous genetic variants are known to contribute to the cardiovascular pathophysiology of FH, several of which have led to the development of approved therapeutic agents. As a positive control to validate our analytical approach, we first performed an association analysis comparing FH subjects with non-FH controls, which successfully identified the *LDLR* p.Trp87Gly variant as being exclusively present in the FH cohort (data not shown). Furthermore, within the CVE-free survival analysis, several variants located in established FH-related genes, including *PCSK9*, *APOB*, and *LDLRAP1*, were identified and served as internal positive controls supporting the validity of the analytical strategy (data not shown).

WES analysis identified a total of 61 variants associated with CVE status in age-stratified HeFH subjects at the exploratory significance threshold of *P* < 1 × 10⁻³ (Table S1). The annotated variant list included synonymous and non-synonymous variants, including missense, nonsense, and frameshift mutations, together with their predicted biological functions and molecular consequences. To prioritize variants with the greatest likelihood of functional relevance, only variants predicted to alter protein structure or expression were retained for further analysis. These included primarily missense, nonsense, and frameshift variants and are presented in Table 2.

**Table 2.** Non-synonymous variants associated with cardiovascular event-free survival past 70 years of age in *LDLR*:c.259T>G (p.Trp87Gly) HeFH.

| Variant<br>(GRCh37/hg19) | Gene | SNP | AF<br>(gnomAD) | Biological/Cellular<br>Function | Consequence | AF<br>(CVE -) | AF<br>(CVE +) | $\chi^2$ | P-value |
| --- | --- | --- | --- | --- | --- | --- | --- | --- | --- |
| 16:70694000C>A | IL-34 | rs4985556 | 0.076 | inflammatory process | stop codon | 0.25 | 0 | 19.25 | 1.15x10<sup>-5</sup> |
| 17:XXXC>A | ECO001 | rs1XXX3 | 0.485 | cell cycle division | missense | 0.7 | 0.3 | 16.49 | 4.90x10<sup>-5</sup> |
| 17:XXXA>G | ECO002 | rs9XXX5 | 0.393 | potassium ion transport | 5'UTR | 0.7 | 0.3 | 16.49 | 4.90x10<sup>-5</sup> |
| X:XXXT>C | ECO006 | rs2XXX4 | 0.453 | transcriptional process | missense | 0.4 | 0.047 | 14.79 | 1.20x10<sup>-4</sup> |
| 17:XXXC>G | ECO007 | rsXXX9 | 0.064 | vascular development | missense | 0.3 | 0.043 | 14.29 | 1.57x10<sup>-4</sup> |
| 2:XXXT>C | ECO016 | rs1XXX9 | 0.051 | wnt signaling pathway | 3' UTR | 0.25 | 0.029 | 12.84 | 3.39x10<sup>-4</sup> |
| 3:XXXA>G | ECO017 | rs6XXX4 | 0.395 | endocytic membrane<br>transport | missense | 0.421 | 0.118 | 12.81 | 3.44x10<sup>-4</sup> |
| X:XXXG>A | ECO018 | rs8XXX0 | 0.417 | lipid transport | 3'UTR | 0.4 | 0.068 | 12.71 | 3.64x10<sup>-4</sup> |
| 20:XXXT>C | ECO020 | rs6XXX1 | 0.346 | receptor-mediated<br>endocytosis | missense | 0.6 | 0.257 | 12.68 | 3.70x10<sup>-4</sup> |
| 9:XXXT>C | ECO021 | rs1XXX2 | 0.330 | nucleocytoplasmic<br>transport | missense | 0.45 | 0.143 | 12.65 | 3.75x10<sup>-4</sup> |
| 16:XXXA>T | ECO027 | rs2XXX8 | 0.169 | ribosome binding | missense | 0.275 | 0.043 | 12.35 | 4.41x10<sup>-4</sup> |
| 7:XXXA>G | ECO037 | rs7XXX4 | 0.119 | lipid metabolism | splice<br>region | 0.35 | 0.086 | 11.95 | 5.46x10<sup>-4</sup> |
| 20:XXXT>A | ECO036 | rs3XXX1 | 0.152 | endothelial cell migration | missense | 0.35 | 0.086 | 11.95 | 5.46x10<sup>-4</sup> |
| 17:XXXA>G | ECO035 | rs8XXX0 | 0.295 | ubiquitin pathway | missense | 0.35 | 0.086 | 11.95 | 5.46x10<sup>-4</sup> |
| 20:47292823C>G | ECO039 | rs6XXX8 | 0.0618 | G protein-coupled<br>receptor signaling | intronic | 0.275 | 0.044 | 11.9 | 5.62x10<sup>-4</sup> |
| 11:XXXG>A | ECO040 | rs1XXX8 | 0.064 | mRNA processing | missense | 0.2 | 0.014 | 11.69 | 6.30x10<sup>-4</sup> |
| 11:XXXC>T | ECO041 | rs7XXX2 | 0.063 | cell proliferation | intronic | 0.2 | 0.014 | 11.69 | 6.30x10<sup>-4</sup> |
| 16:XXXG>A | ECO046 | rs2XXX2 | 0.399 | receptor-mediated<br>endocytosis | splice<br>region | 0.5 | 0.191 | 11.32 | 7.67x10<sup>-4</sup> |
| 22:XXXA>G | ECO045 | rs1XXX0 | 0.140 | mRNA processing | missense | 0.2 | 0.015 | 11.32 | 7.67x10<sup>-4</sup> |
| 17:XXXG>T | ECO048 | rs6XXX5 | 0.416 | receptor guanylyl<br>cyclase signaling | missense | 0.525 | 0.214 | 11.16 | 8.35x10<sup>-4</sup> |
| 2:XXXC>G | ECO051 | rs3XXX3 | 0.024 | G protein-coupled<br>receptor signaling | missense | 0.15 | 0 | 11.11 | 8.61x10<sup>-4</sup> |
| 19:XXXC>T | ECO053 | rs3XXX4 | 0.179 | receptor-mediated<br>endocytosis | missense | 0.15 | 0 | 11.11 | 8.61x10<sup>-4</sup> |
| 12:XXXT>C | ECO049 | rs1XXX0 | 0.023 | mannosyltransferase<br>activity | intronic | 0.15 | 0 | 11.11 | 8.61x10<sup>-4</sup> |
| 7:XXXC>T | ECO058 | rs3XXX6 | 0.023 | lipid metabolism | intronic | 0.225 | 0.029 | 10.91 | 9.55x10<sup>-4</sup> |

Table includes all non-synonymous variant associations with a p-value <0.001 and an odds ratio (relative odds of no CVE) >1.
Biological/Cellular function terms extracted from Gene Ontology annotations. With the exception of *IL34*, all identified candidate genes were anonymized using ECO acronyms to preserve confidentiality in anticipation of potential intellectual property and patent protection.
Abbreviations: CVE, cardiovascular events (“-” patients with no events, “+” patients with events); AF, allele frequency; $\chi^2$ , Chi-square

All chromosomal positions were mapped using the *Homo sapiens* reference genome assembly GRCh37 (hg19), and associations were evaluated by comparing allele frequencies between the CVE(-) and CVE(+) groups using Pearson’s χ² test.

Among the prioritized variants, a stop-gain variant resulting from the SNP rs4985556 in the *IL34* gene emerged as the strongest association signal. This variant introduces a premature stop codon in IL-34 (c.639C>A; p.Tyr213*) and was significantly enriched among CVE-free survivors (AF in CVE(-) = 0.25 vs. AF in CVE(+) = 0.00; χ² = 19.25; *P* = 1.15 × 10⁻⁵) (Table 2; Figure S1).

The predicted biological functions of all prioritized genes are summarized in Figure S2. To further evaluate the contribution of these variants to CVE-free survival, Firth logistic regression analyses were performed on all candidate variants presented in Table 2. Twelve variants remained significantly associated with CVE-free survival, with ORs ranging from 4.2 to 31.8 and *P* values reaching <1 × 10⁻⁴.

The strongest association was again observed for the *IL34* stop-gain variant (rs4985556; c.639C>A; p.Tyr213*), which was associated with a markedly increased likelihood of CVE-free survival (OR = 20.9; 95% CI: 2.7–2841.7; *P* = 3.45 × 10⁻⁴) (Figure 2).

**Figure 2.**
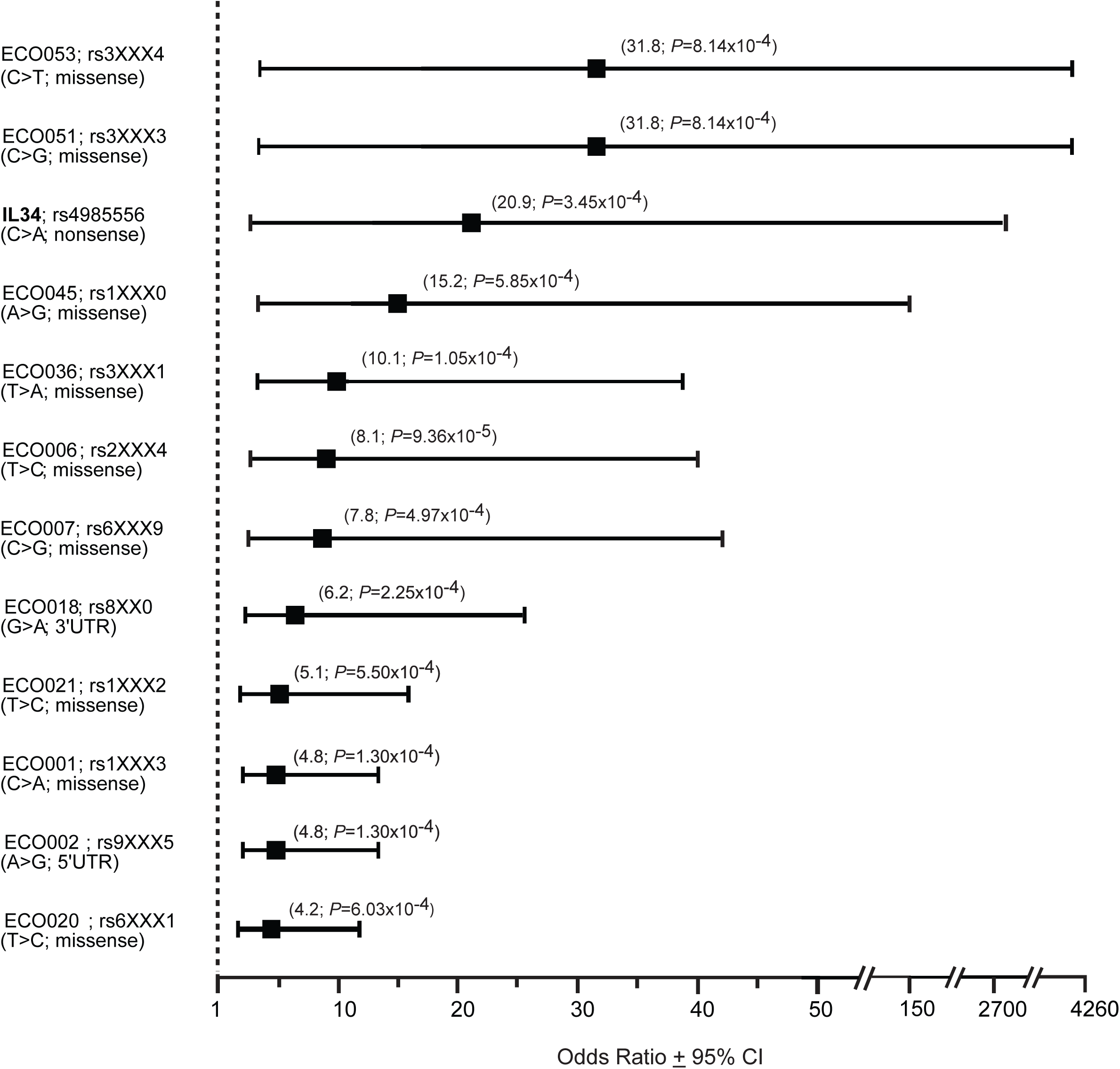
Association of candidate genetic variants with cardiovascular event-free survival in familial hypercholesterolemia. Forest plot showing the odds ratios (ORs) and 95% confidence intervals derived from Firth logistic regression analyses evaluating the association between nonsynonymous genetic variants and cardiovascular event-free survival [CVE(-)] in FH patients. Only variants meeting the predefined selection criteria (*P* < 1 × 10^-3^ and OR > 1) are presented. All displayed variants showed significant associations with CVE(-) following Firth logistic regression analysis.

With the exception of *IL34*, all identified candidate genes were anonymized using ECO acronyms to preserve confidentiality in anticipation of potential intellectual property and patent protection.

## Discussion

This study identifies a significant association between a loss-of-function variant in *IL34* and CVE-free survival among French-Canadian HeFH patients exposed to lifelong elevations in LDL-C levels. These findings extend our previous work identifying non-genetic markers of CVE-free survival in FH, including female sex, higher HDL-C, elevated adiponectin, and non-smoking status (20). Together, these observations support the concept that cardiovascular resilience in FH is likely multifactorial, reflecting the combined influence of genetic variation, lipid-related factors, inflammation, metabolic profile, environmental exposures and sex-specific biology.

IL-34 is a secreted homodimeric cytokine expressed in multiple tissues, including the brain, lungs, lymph nodes, liver, heart, and vascular endothelium. IL-34 was identified as a second ligand of the colony-stimulating factor 1 receptor (CSF-1R), alongside macrophage colony-stimulating factor (M-CSF). Activation of CSF-1R signaling by M-CSF is known to promote inflammatory processes involved in atherogenesis. Although the respective downstream signaling pathways activated by IL-34 and M-CSF remain incompletely characterized, both cytokines stimulate monocyte differentiation into mature macrophages (23–25). Consequently, IL-34 may contribute to atherosclerotic disease progression through mechanisms similar to those described for M-CSF.

Supporting this hypothesis, Liu et al. demonstrated that IL-34 promotes macrophage foam cell formation through CSF-1R-mediated activation of the MAPK/p38 pathway, resulting in increased CD36 expression, enhanced oxidized LDL uptake, and elevated production of pro-inflammatory cytokines including IL-6, TNF-α, and IL-1β (26). These pathways are well-established contributors to vascular inflammation and atherosclerotic plaque development.

Cardiovascular disease is now recognized as arising from the interplay between dyslipidemia, inflammation, and endothelial dysfunction, each of which contributes to the initiation and progression of atherosclerosis (27, 28). The present findings suggest that genetic variation affecting IL-34 signaling may modulate the inflammatory component of atherosclerotic disease and partially offset the adverse effects of lifelong exposure to elevated LDL-C levels in FH. Not to mention that these findings also extend our previous work identifying non-genetic markers of CVE-free survival in FH (20). Together, these observations support the concept that cardiovascular resilience in FH could likely be multifactorial, reflecting the combined influence of genetic variation, lipid-related factors, inflammation, metabolic profile, environmental exposures and sex-specific biology.

The *IL34* variant identified in this study (rs4985556; c.639C>A; p.Tyr213*) is a nonsense variant located within the final exon of the gene and is predicted to truncate the encoded protein by approximately 12%. Because nonsense-mediated decay is generally less efficient for variants located in terminal exons, the resilience (event-free) effect observed here may not be solely attributable to reduced gene expression. Alternative mechanisms could include altered protein structure, impaired homodimerization, reduced receptor binding affinity, or modification of downstream CSF-1R signaling. These hypotheses warrant future functional investigation.

A proposed mechanistic model is presented in Figure 3. Under physiological conditions, IL-34 and/or CSF-1 bind to CSF-1R on macrophages, inducing receptor dimerization and activation of downstream signaling pathways including RAS–RAF–MEK–ERK and PI3K–Akt. These pathways promote macrophage proliferation, differentiation, survival, and foam-cell formation, thereby contributing to vascular inflammation and plaque development. We hypothesize that the *IL34* loss-of-function variant may attenuate CSF-1R signaling, ultimately reducing macrophage activation and foam-cell formation and thereby decreasing the risk of CVE.

**Figure 3.**
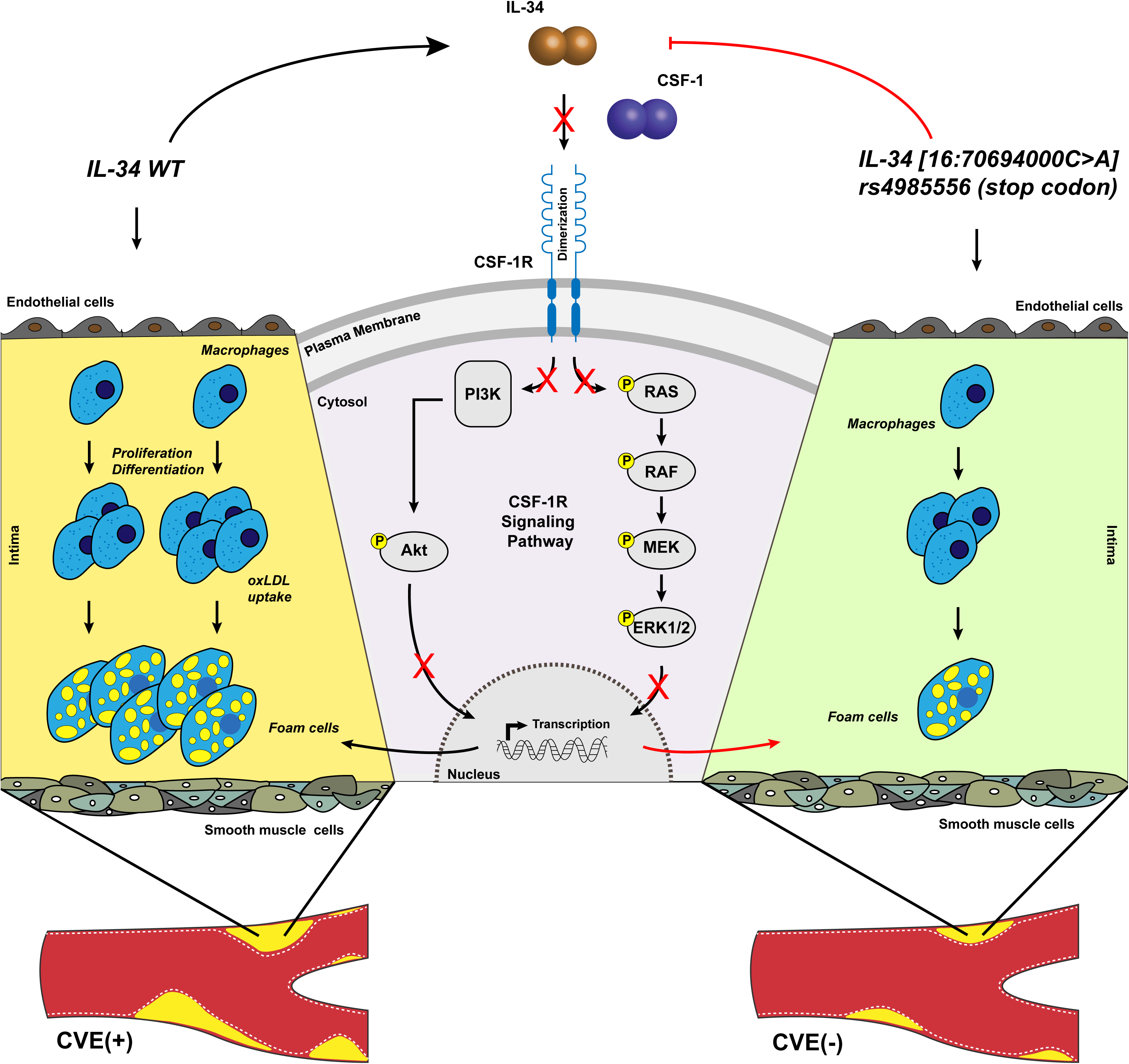
Proposed mechanism underlying the cardiovascular event-free role of the *IL34* loss-of-function variant (rs4985556; c.639C>A; p.Tyr213). IL-34 acts through modulation of colony-stimulating factor 1 receptor (CSF-1R) signaling in macrophages.*\** Under physiological conditions, IL-34 binding to CSF-1R promotes macrophage proliferation, differentiation, and survival, thereby contributing to vascular inflammation, foam-cell formation, and atherosclerotic plaque development. The *IL34* loss-of-function variant (rs4985556; p.Tyr213*) is hypothesized to attenuate CSF-1R signaling, resulting in reduced macrophage activation and foam-cell formation, ultimately limiting plaque progression within the arterial wall and contributing to cardiovascular event-free survival.

The growing evidence linking IL-34 to cardiovascular disease further supports the biological plausibility of our findings. Elevated circulating IL-34 concentrations have been associated with acute ischemic stroke (29), acute myocardial infarction (30), chronic heart failure (31), coronary artery disease (32), and ischemic cardiomyopathy severity (33). Our findings provide complementary human genetic evidence supporting a potential role for IL-34 in atherosclerotic cardiovascular disease. These findings are also consistent with growing evidence that modulation of interleukin-mediated inflammation may reduce cardiovascular risk independently of lipid lowering. Landmark studies targeting the IL-1β signaling cascade (CANTOS) and broader immunity pathways with low-dose colchicine (COLCOT and LoDoCo2) have demonstrated that suppression of vascular inflammation lowers the incidence of major CVE (34). More recently, inhibition of IL-6 signaling with ziltivekimab has shown promising anti-inflammatory effects and is currently being evaluated in large cardiovascular outcome trials (35). Together, these observations reinforce the concept that multiple cytokine pathways, including the IL-34/CSF-1R axis identified in the present study, may represent attractive therapeutic targets for cardiovascular prevention.

From a translational perspective, these results suggest that IL-34 may represent a novel therapeutic target for reducing cardiovascular risk through modulation of inflammatory pathways independent of lipid lowering. Beyond *IL34*, eleven additional variants were significantly associated with CVE-free survival and warrant further investigation. These variants are located in genes encoding proteins involved in diverse biological processes relevant to atherosclerosis and cardiovascular disease, including inflammatory signaling, vascular development and endothelial cell migration, lipid metabolism and transport, receptor-mediated endocytosis, G protein-coupled receptor and Wnt signaling, potassium ion transport, cell-cycle regulation and proliferation, transcriptional and post-transcriptional regulation, nucleocytoplasmic transport, ubiquitin-mediated protein turnover, and ribosomal function (Supp. Fig 2). Together, these findings highlight multiple biological pathways that may contribute to cardiovascular resilience and represent potential targets for future mechanistic and therapeutic studies.

Several limitations should be acknowledged. First, the study was conducted in a relatively small number of subjects selected for WES. Second, the significance threshold used for variant prioritization was exploratory and does not correspond to conventional genome-wide significance thresholds. However, the study leveraged a highly homogeneous founder population from Saguenay–Lac-Saint-Jean, a setting in which rare variants of relatively large effect may be enriched and more readily detected. Similar founder-population approaches have previously led to the identification of clinically relevant therapeutic targets, involving the *APOC3, ANGPTL3, ANGPTL8, PCSK9* pathways, among others, which ultimately contributed to the development of novel therapies for unmet medical needs (36–39).

Finally, independent validation in external FH cohorts and large population-based biobanks will be required to confirm the associations reported here. Functional characterization of prioritized variants, including protein quantification, receptor signaling studies, and cellular assays, will also be necessary to establish their biological relevance. We do also acknowledge the possibility of having residual confounding factors that might influence the CVE trajectory, in addition to the previously identified clinical, biological and environmental resilience markers (20), however, given the small sample size, multivariate adjustment may be limited. Importantly, these genetic findings complement our previous work identifying biological and environmental factors associated with unexpected CVE-free survival among FH patients (20). Collectively, these observations further support the concept that FH, including HoFH, may serve as a unique human model for the identification of genetic, biological, and environmental determinants of unexpected survival despite a profound lifelong predisposition to atherosclerotic cardiovascular disease and premature mortality.

## Conclusion

This study investigated genetic determinants associated with event-free survival in HeFH phenotypes, comparing patients aged ≥70 years without reported CVE [CVE(-)] with younger patients who had experienced premature CVE [CVE(+)]. The premature stop-gain variant rs4985556 in the *IL34* gene (c.639C>A; p.Tyr213*) was observed exclusively among CVE(-) patients and was associated with markedly increased odds of CVE-free survival.

These findings identify *IL34* as a potential marker of event-free survival and support its role as a promising candidate for further investigation in FH, a condition characterized by markedly elevated lifetime risk of atherosclerotic cardiovascular disease and early death without treatment. Therapeutic strategies aimed at modulating IL-34 activity, including RNA-based or gene-targeted approaches, may represent an interesting avenue for future research.

In addition to *IL34*, 11 other variants were strongly associated with the CVE(-) phenotype in FH, with ORs exceeding 10-fold for several candidates. These variants warrant further investigation to elucidate their mechanisms of action, effects on cellular signaling, and potential biological roles in cardiovascular protection beyond familial hypercholesteremia.

## Data Availability

Data can be provided upon request

## Supplementary Figures Legends

**Figure S1.** Manhattan plot showing exome-wide associations between genetic variants and cardiovascular event status in age-stratified heterozygous familial hypercholesterolemia (HeFH) patients carrying the *LDLR* c.259T>G (p.Trp87Gly) variant. Variants reaching the exploratory significance threshold (*P* < 1 × 10^-3^) were selected as candidates for further analyses. Annotated variants (ECO001–ECO008), including *IL34*, represent nonsynonymous variants significantly associated with cardiovascular event-free survival [CVE(-)] or cardiovascular events [CVE(+)] and were prioritized for downstream validation analyses.

**Figure S2.** Circular plot summarizing the predicted biological and cellular functions of genes associated with cardiovascular event status in age-stratified heterozygous familial hypercholesterolemia (HeFH) patients carrying the *LDLR* c.259T>G (p.Trp87Gly) variant. Functional annotations were derived from Gene Ontology (GO) and related biological databases and were used to classify candidate genes according to their putative roles in cellular signaling, inflammation, lipid metabolism, vascular biology, and other processes relevant to cardiovascular disease.

## References

1. Tokgozoglu L, Kayikcioglu M. Familial Hypercholesterolemia: Global Burden and Approaches. Curr Cardiol Rep. 2021;23(10):151.

2. Aljenedil S, Ruel I, Watters K, Genest J. Severe xanthomatosis in heterozygous familial hypercholesterolemia. J Clin Lipidol. 2018;12(4):872–7.

3. Watts GF, Gidding SS, Mata P, Pang J, Sullivan DR, Yamashita S, et al. Familial hypercholesterolaemia: evolving knowledge for designing adaptive models of care. Nat Rev Cardiol. 2020;17(6):360–77.

4. Ahmad Z, Agarwala A, Cuchel M, Barton Duell P, Hegele RA, Hudgins L, et al. Update on familial hypercholesterolemia: An expert clinical consensus from the National Lipid Association. J Clin Lipidol. 2026;20(4):708–37.

5. Henderson R, O’Kane M, McGilligan V, Watterson S. The genetics and screening of familial hypercholesterolaemia. J Biomed Sci. 2016;23:39.

6. Hu P, Dharmayat KI, Stevens CAT, Sharabiani MTA, Jones RS, Watts GF, et al. Prevalence of Familial Hypercholesterolemia Among the General Population and Patients With Atherosclerotic Cardiovascular Disease: A Systematic Review and Meta-Analysis. Circulation. 2020;141(22):1742–59.

7. Bchetnia M, Bouchard L, Mathieu J, Campeau PM, Morin C, Brisson D, et al. Genetic burden linked to founder effects in Saguenay-Lac-Saint-Jean illustrates the importance of genetic screening test availability. J Med Genet. 2021;58(10):653–65.

8. Gaudet D, Vohl MC, Perron P, Tremblay G, Gagne C, Lesiege D, et al. Relationships of abdominal obesity and hyperinsulinemia to angiographically assessed coronary artery disease in men with known mutations in the LDL receptor gene. Circulation. 1998;97(9):871–7.

9. Schunkert H, Natarajan P, Samani NJ. The Inherited Basis of Coronary Artery Disease. N Engl J Med. 2026;394(6):576–87.

10. Yu Y, Chen L, Zhang H, Fu Z, Liu Q, Zhao H, et al. Association Between Familial Hypercholesterolemia and Risk of Cardiovascular Events and Death in Different Cohorts: A Meta-Analysis of 1.1 Million Subjects. Front Cardiovasc Med. 2022;9:860196.

11. Conchinha A, Rodrigues A, Pack T, Cunha S, Santos A. Familial Hypercholesterolaemia and the Risk of Cardiovascular Events. Cureus. 2025;17(11):e97656.

12. Moorjani S, Roy M, Torres A, Betard C, Gagne C, Lambert M, et al. Mutations of low-density-lipoprotein-receptor gene, variation in plasma cholesterol, and expression of coronary heart disease in homozygous familial hypercholesterolaemia. Lancet. 1993;341(8856):1303–6.

13. Nordestgaard BG, Chapman MJ, Humphries SE, Ginsberg HN, Masana L, Descamps OS, et al. Familial hypercholesterolaemia is underdiagnosed and undertreated in the general population: guidance for clinicians to prevent coronary heart disease: consensus statement of the European Atherosclerosis Society. Eur Heart J. 2013;34(45):3478–90a.

14. Tromp TR, Hartgers ML, Hovingh GK, Vallejo-Vaz AJ, Ray KK, Soran H, et al. Worldwide experience of homozygous familial hypercholesterolaemia: retrospective cohort study. Lancet. 2022;399(10326):719–28.

15. Mulder J, Reijman MD, Kusters DM, Boersma E, Alnouri F, Blom DJ, et al. Homozygous Familial Hypercholesterolemia Is a Life-Limiting Condition: Medical Life-Trajectories in the Post-2010 Era. J Am Coll Cardiol. 2025;85(19):1898–903.

16. Patel J, Shah S, Reddy A, Prajapati D, Pandya A, Sawhney S. Novel Approaches to Lipid Management: Beyond Statins and PCSK9 Inhibitors. J Clin Med Res. 2026;18(3):121–41.

17. Jianu N, Nitu ET, Merlan C, Nour A, Buda S, Suciu M, et al. A Comprehensive Review of the Latest Approaches to Managing Hypercholesterolemia: A Comparative Analysis of Conventional and Novel Treatments: Part II. Pharmaceuticals (Basel). 2025;18(8).

18. Low-Kam C, Rhainds D, Lo KS, Provost S, Mongrain I, Dubois A, et al. Whole-genome sequencing in French Canadians from Quebec. Hum Genet. 2016;135(11):1213–21.

19. Cruz Marino T, Leblanc J, Pratte A, Tardif J, Thomas MJ, Fortin CA, et al. Portrait of autosomal recessive diseases in the French-Canadian founder population of Saguenay-Lac-Saint-Jean. Am J Med Genet A. 2023;191(5):1145–63.

20. Khoury E, Brisson D, Roy N, Tremblay G, Gaudet D. Identifying Markers of Cardiovascular Event-Free Survival in Familial Hypercholesterolemia. J Clin Med. 2020;10(1).

21. Randomised trial of cholesterol lowering in 4444 patients with coronary heart disease: the Scandinavian Simvastatin Survival Study (4S). Lancet. 1994;344(8934):1383–9.

22. Li H. Aligning sequence reads, clone sequences and assembly contigs with BWA-MEM. arXiv preprint arXiv:13033997. 2013.

23. Nakamichi Y, Udagawa N, Takahashi N. IL-34 and CSF-1: similarities and differences. J Bone Miner Metab. 2013;31(5):486–95.

24. Munoz-Garcia J, Cochonneau D, Teletchea S, Moranton E, Lanoe D, Brion R, et al. The twin cytokines interleukin-34 and CSF-1: masterful conductors of macrophage homeostasis. Theranostics. 2021;11(4):1568–93.

25. Baghdadi M, Umeyama Y, Hama N, Kobayashi T, Han N, Wada H, et al. Interleukin-34, a comprehensive review. J Leukoc Biol. 2018;104(5):931–51.

26. Liu Q, Fan J, Bai J, Peng L, Zhang T, Deng L, et al. IL-34 promotes foam cell formation by enhancing CD36 expression through p38 MAPK pathway. Sci Rep. 2018;8(1):17347.

27. Medina-Leyte DJ, Zepeda-Garcia O, Dominguez-Perez M, Gonzalez-Garrido A, Villarreal-Molina T, Jacobo-Albavera L. Endothelial Dysfunction, Inflammation and Coronary Artery Disease: Potential Biomarkers and Promising Therapeutical Approaches. Int J Mol Sci. 2021;22(8).

28. Members WC, Blumenthal RS, Morris PB, Gaudino M, Johnson HM, Anderson TS, et al. 2026 ACC/AHA/AACVPR/ABC/ACPM/ADA/AGS/APhA/ASPC/NLA/PCNA Guideline on the Management of Dyslipidemia: A Report of the American College of Cardiology/American Heart Association Joint Committee on Clinical Practice Guidelines. Circulation.0(0).

29. Huang X, Li F, Yang T, Li H, Liu T, Wang Y, et al. Increased serum interleukin-34 levels as a novel diagnostic and prognostic biomarker in patients with acute ischemic stroke. J Neuroimmunol. 2021;358:577652.

30. Fan Q, Tao R, Zhang H, Xie H, Xi R, Wang F, et al. Interleukin-34 Levels Were Associated with Prognosis in Patients with Acute Myocardial Infarction. Int Heart J. 2019;60(6):1259–67.

31. Tao R, Fan Q, Zhang H, Xie H, Lu L, Gu G, et al. Prognostic Significance of Interleukin-34 (IL-34) in Patients With Chronic Heart Failure With or Without Renal Insufficiency. J Am Heart Assoc. 2017;6(4).

32. Li Z, Jin D, Wu Y, Zhang K, Hu P, Cao X, et al. Increased serum interleukin-34 in patients with coronary artery disease. J Int Med Res. 2012;40(5):1866–70.

33. Xi R, Fan Q, Yan X, Zhang H, Xie H, Gu G, et al. Increased Serum Interleukin-34 Levels Are Related to the Presence and Severity of Cardiac Dysfunction in Patients With Ischemic Cardiomyopathy. Front Physiol. 2018;9:904.

34. Ridker PM, Everett BM, Thuren T, MacFadyen JG, Chang WH, Ballantyne C, et al. Antiinflammatory Therapy with Canakinumab for Atherosclerotic Disease. N Engl J Med. 2017;377(12):1119–31.

35. Ridker PM, Devalaraja M, Baeres FMM, Engelmann MDM, Hovingh GK, Ivkovic M, et al. IL-6 inhibition with ziltivekimab in patients at high atherosclerotic risk (RESCUE): a double-blind, randomised, placebo-controlled, phase 2 trial. Lancet. 2021;397(10289):2060–9.

36. Gaudet D, Brisson D, Tremblay K, Alexander VJ, Singleton W, Hughes SG, et al. Targeting APOC3 in the familial chylomicronemia syndrome. N Engl J Med. 2014;371(23):2200–6.

37. Abadi A, Chen YQ, Khalid S, Williams K, Zhen EY, Li H, et al. Unraveling the physiological impact of ANGPTL8 loss-of-function variants in humans. Sci Rep. 2026.

38. Abifadel M, Varret M, Rabes JP, Allard D, Ouguerram K, Devillers M, et al. Mutations in PCSK9 cause autosomal dominant hypercholesterolemia. Nat Genet. 2003;34(2):154–6.

39. Musunuru K, Pirruccello JP, Do R, Peloso GM, Guiducci C, Sougnez C, et al. Exome sequencing, ANGPTL3 mutations, and familial combined hypolipidemia. N Engl J Med. 2010;363(23):2220–7.

